# DeepSARS: simultaneous diagnostic detection and genomic surveillance of SARS-CoV-2

**DOI:** 10.1101/2021.08.16.21262126

**Authors:** Alexander Yermanos, Kai-Lin Hong, Andreas Agrafiotis, Jiami Han, Sarah Nadeau, Cecilia Valenzuela, Asli Azizoglu, Roy Ehling, Beichen Gao, Michael Spahr, Daniel Neumeier, Ching-Hsiang Chang, Andreas Dounas, Ezequiel Petrillo, Ina Nissen, Elodie Burcklen, Mirjam Feldkamp, Christian Beisel, Annette Oxenius, Miodrag Savic, Tanja Stadler, Fabian Rudolf, Sai T. Reddy

## Abstract

The continued spread of SARS-CoV-2 and emergence of new variants with higher transmission rates and/or partial resistance to vaccines has further highlighted the need for large-scale testing and genomic surveillance. However, current diagnostic testing (e.g., PCR) and genomic surveillance methods (e.g., whole genome sequencing) are performed separately, thus limiting the detection and tracing of SARS-CoV-2 and emerging variants. Here, we developed DeepSARS, a high-throughput platform for simultaneous diagnostic detection and genomic surveillance of SARS-CoV-2 by the integration of molecular barcoding, targeted deep sequencing, and computational phylogenetics. DeepSARS enables highly sensitive viral detection, while also capturing genomic diversity and viral evolution. We show that DeepSARS can be rapidly adapted for identification of emerging variants, such as alpha, beta, gamma, and delta strains, and profile mutational changes at the population level. DeepSARS sets the foundation for quantitative diagnostics that capture viral evolution and diversity.

Graphical abstract
DeepSARS uses molecular barcodes (BCs) and multiplexed targeted deep sequencing (NGS) to enable simultaneous diagnostic detection and genomic surveillance of SARS-CoV-2.

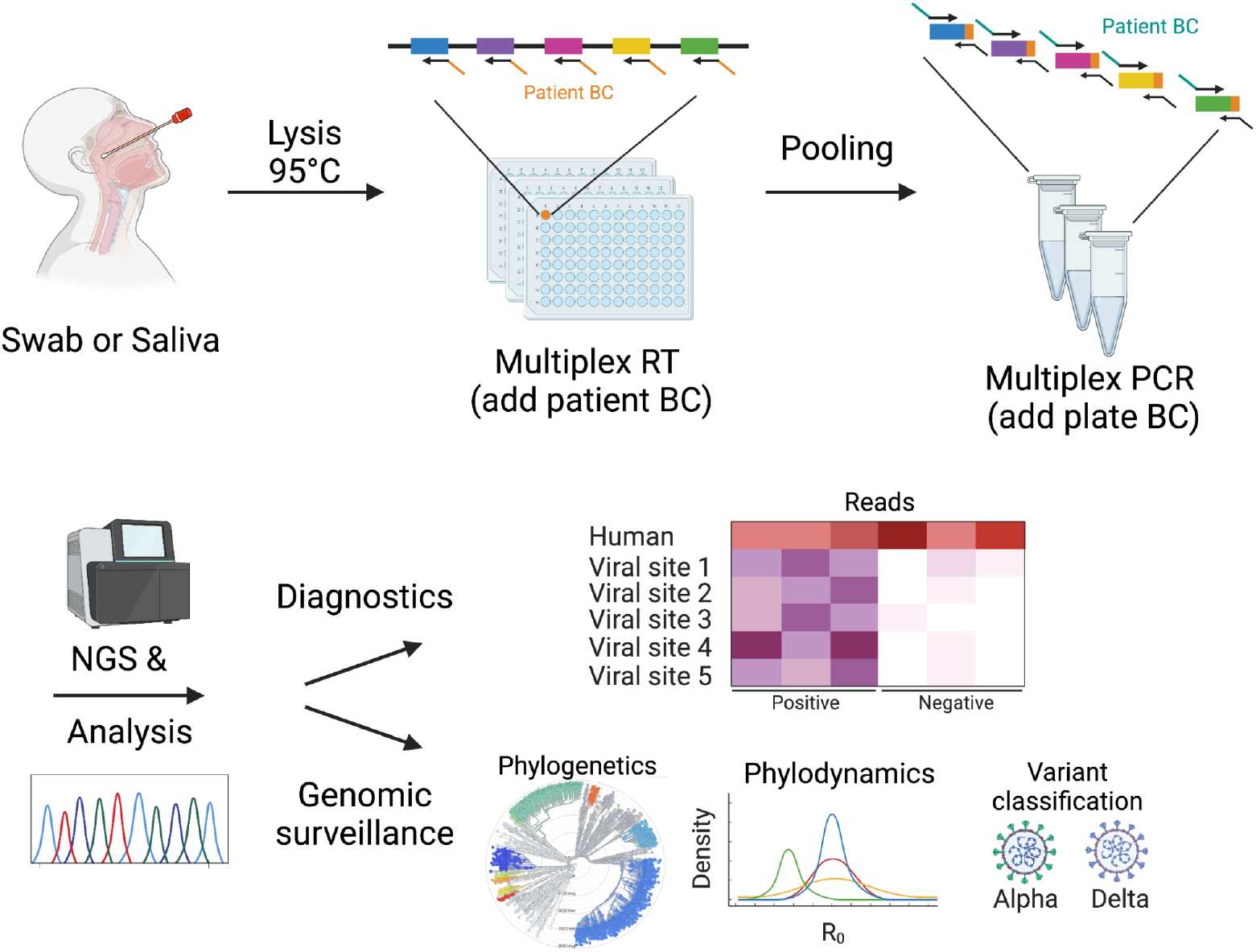

## Introduction

The coronavirus disease 2019 (COVID-19) pandemic, caused by severe acute respiratory syndrome coronavirus 2 (SARS-CoV-2), has presented a global health challenge of a magnitude not experienced in over a century. In response to the pandemic, large-scale diagnostic capabilities have been implemented to track viral spread and inform public health strategies such as contact tracing and social distancing (Shental et al. 2020; Hogan, Sahoo, and Pinsky 2020; Barak et al. 2021; Mercer and Salit 2021). The current tests for COVID-19 rely on reverse transcription (RT) and PCR test or rapid antigen detection assays, which are used for detection of SARS-CoV-2 nucleic acids or proteins, respectively. These diagnostics, paired with vaccination, have greatly succeeded in reducing both the prevalence and severity of COVID-19 in some regions of the world. However, the ongoing evolution of SARS-CoV-2 and the emergence of variants of concern (VoC) such as alpha (B.1.1.7), beta (B.1.351), gamma (P.1), and delta (B.1.617.2) that have higher transmission rates and/or possess partial resistance to vaccines and antibody therapeutics threaten to alter the trajectory of the pandemic (Wang et al. 2021; Hoffmann et al. 2021; Wibmer et al. 2021; Garcia-Beltran et al. 2021; McCormick, Jacobs, and Mellors 2021; Harvey et al. 2021; Planas et al. 2021).

Genomic surveillance is crucial to monitoring the continuously mutating SARS-CoV-2 and preventing wide-spread circulation of current VoC, as well as future variants with potentially enhanced vaccine- and antibody-resistance. Genomic surveillance is most often performed by whole genome sequencing (WGS) of the ∼30,000 base genome of SARS-CoV-2, which consists of RNA-extraction (typically from nasopharyngeal swab samples) followed by molecular library preparation and deep sequencing. In addition to variant classification, WGS provides a powerful tool to infer evolutionary parameters such as mutation rates, effective reproductive number (R_e_), and genetic drift; and the implementation of WGS combined with large-scale databases (e.g., GISAID) is providing unprecedented insight on viral evolution in real-time and across geographic regions (Scire, Huisman, and Stadler 2021; Hodcroft et al. 2021; Müller et al. 2021; Popa et al. 2020). Although major limitations of WGS are that it is often resource-, cost-, and time-intensive, and while scale-up of WGS has led globally to more than 2 million sequenced genomes (as of June 2021 on GISAID), this pales in comparison to the number of infections globally (∼183 million as of June 2021). Furthermore, WGS is unevenly applied across countries, for example in the United Kingdom 10.9% of positive patients have been sequenced, whereas, in countries such as Brazil and South Africa only 0.11% and 0.52% have been sequenced, respectively (as of June 2021). This problem becomes even more critical considering many of the countries with the lowest fraction of sequenced genomes are also the ones where infection rates are highest, and thus represent hotspots for viral evolution and potential origins for future VoC. Therefore, increasing the throughput and capacity for genomic surveillance of SARS-CoV-2 is essential for monitoring and preventing the spread of existing and emerging VoC.

While PCR tests have achieved large-scale implementation for SARS-CoV-2 detection and diagnosis, they are unable to produce information on viral sequence diversity and evolution; in contrast while, WGS does enable genomic surveillance and detection of variants, the costs and resources required limit its implementation at large-scale globally. Targeted amplification combined with deep sequencing offers a compromise between these two methods, as demonstrated by several recent studies that have described protocols for SARS-CoV-2 testing (Aynaud et al. 2021; Bhoyar et al. 2021; Bloom et al. 2020; Yelagandula et al. 2021). However, these methods currently rely upon selectively sequencing only a few conserved regions of the SARS-CoV-2 genome. For example, the SARSseq method described by Yelagandula et al., relies on only sequencing two short regions (∼ 70 bp) of the N gene (Yelagandula et al. 2021); similarly, Bloom et al. developed SwabSeq which targets only a single, minimal region (26 bp) of the *S* gene of SARS-CoV-2 (Bloom et al. 2021). Aynaud et al. established the C19-SPAR-Seq method for targeted sequencing of five regions across the viral genome, however, only one of them corresponds to the spike protein’s receptor binding domain (RBD) (Aynaud et al. 2021), where characteristic mutations associated with VoC are found (Harvey et al. 2021). While the diagnostic potential of these methods has been extensively explored, the minimal amount of sequence-space captured excludes their use for genomic surveillance applications such as evolutionary inference and VoC classification.

Here, we describe DeepSARS, a high-throughput platform that can simultaneously perform diagnostic detection and genomic surveillance of SARS-CoV-2. By combining patient sample-specific molecular barcoding, targeted and multiplexed amplicon deep sequencing and computational phylogenetics, DeepSARS provides highly sensitive and specific detection of virus, while also capturing genomic diversity in highly variable regions of the SARS-CoV-2 genome. We initially developed and benchmarked the diagnostic capability of DeepSARS using synthetic RNA templates of SARS-CoV-2, which resulted in viral detection with as low as 10 copies of virus per sample. Next, we applied DeepSARS on human samples, including both nasopharyngeal swabs and saliva samples from COVID-19-positive and -negative individuals, which demonstrated detection sensitivities and specificities similar to the current standard of PCR tests. Importantly, DeepSARS was also able to recover sequence information for approximately 20% of the full genome, which enabled the inference of phylodynamic parameters such as the effective reproductive number and temporal dynamics of local outbreaks. Finally, we demonstrated the ability to rapidly incorporate novel viral sites into the DeepSARS framework to classify VoCs, such as alpha, beta, gamma, and delta strains, and could further detect how individual mutations change over time on a population level.

## RESULTS

### Design of a molecular barcoding strategy to link patient-specific information with deep sequencing

DeepSARS uses an integrated pipeline of experimental library preparation, deep sequencing and computational phylogenetic analysis for the simultaneous detection and genomic surveillance of SARS-CoV-2. The experimental workflow is based on the stepwise incorporation of molecular barcodes for the tagging of RNA viral genes across multiple 96-well plates (Figure 1A). Each well on a 96-well plate corresponds to a single patient sample and molecular barcodes (patientBC) are added during the initial reverse-transcription (RT) step of the viral genome, thereby allowing sequence-based patient identification. The first 96 patientBC sequences were designed to be 10 base pairs (bp) in length; and in order to avoid patient misclassification, sequence diversity was designed to maximize the number of mutations between any two patientBCs on a given plate (Figure S1). Importantly, the addition of patientBCs during the RT step enables pooling of all patient samples from a 96-well plate, whereby a second plate sequence-barcode (plateBC) is added by PCR via annealing to overhangs introduced during the RT step. This pooling step simplifies the workflow and supports sample preparation and parallelization of many 96-well plates. Moreover, the two sequence-barcodes ensure the unambiguous identification of both the origin well (patient) and 96-well plate. A multiplexed set of forward primers targeting the viral regions of interest (and Illumina sequencing adaptors) enables PCR amplification of libraries for deep sequencing.

**Figure 1.**
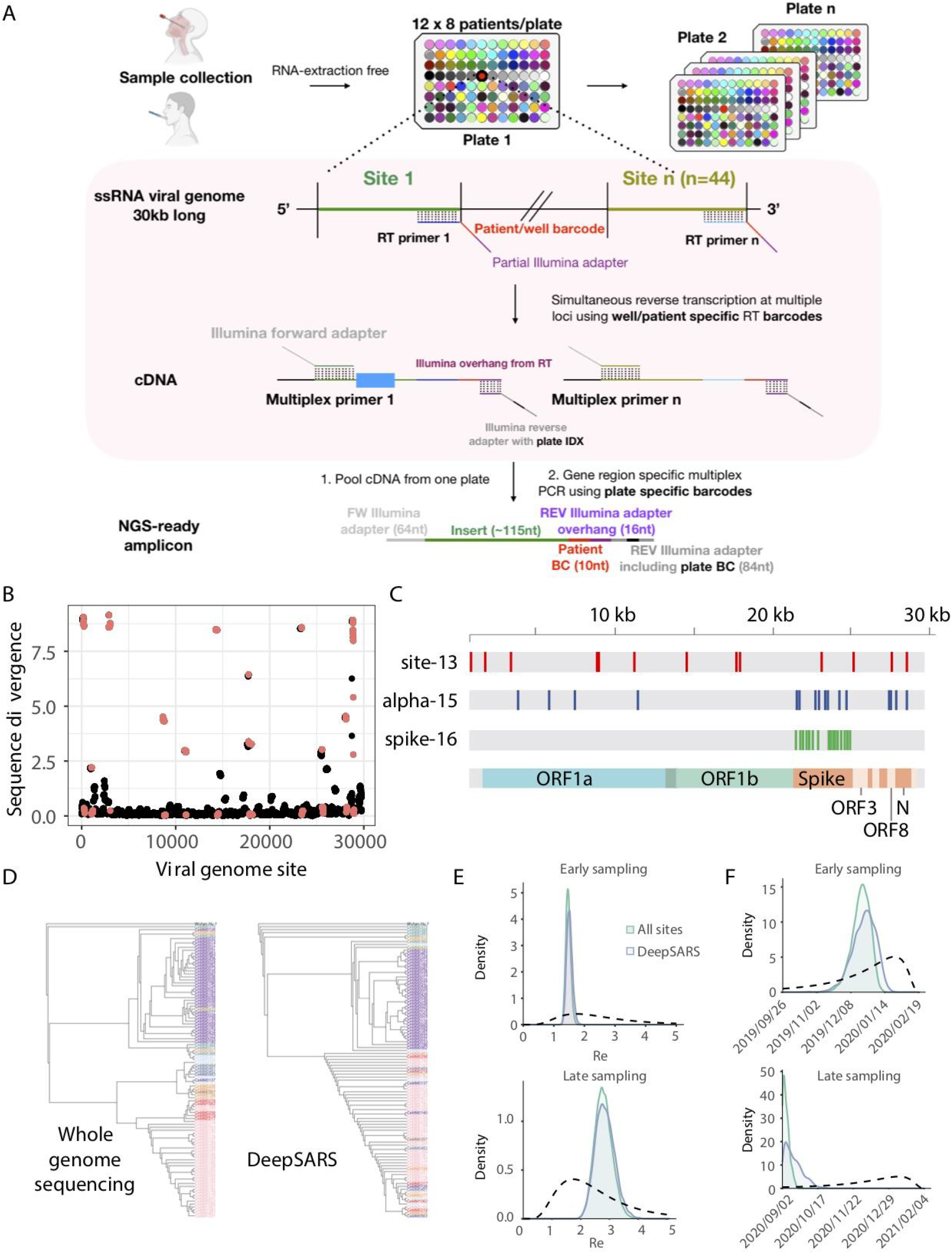
DeepSARS enables simultaneous diagnostic testing and genomic surveillance of SARS-CoV-2. **A**. DeepSARS workflow consists of patient sample collection from nasopharyngeal swab or saliva (without RNA extraction) followed by targeted and multiplexed library preparation for deep sequencing. The multiplexing strategy uses well-specific primers to introduce a patient-barcode during reverse transcription. Samples are then pooled and multiplexed PCR is performed with a plate-specific barcode. **B**. SARS-CoV-2 sequence diversity for each nucleotide based on whole genome sequencing data from 2,825 samples collected between December 2019 and March 2020. Red points indicate regions covered by the site-13 primer set in DeepSARS. **C**. Colored lines represent sites of the SARS-CoV-2 genome targeted by the three primer sets of DeepSARS tested in this study: site-13, alpha-15 and spike-16. **D**. Maximum likelihood trees generated on 100 samples from previous study that profiled an Austrian outbreak (Popa et al. 2020); trees are inferred using either WGS or the sites targeted by DeepSARS. Color corresponds to variant classification (based on geographic location) and tip name corresponds to the individual sequence. Tree was rooted using a reference sequence recovered from Wuhan (NCBI: MN908947). **E**. Phylodynamic estimates of the effective reproductive number (R_e_) using either WGS or the sites covered by DeepSARS. For each tree, 30 sequences were sampled from Italy at two different time points (prior to March 8, 2020) or at a later time point where the majority of sequences correspond to the alpha variant (between February 1, 2021 to March 15, 2021). Dotted line indicates the prior distribution. **F**. Phylodynamic estimates using the same sequences in **E** but depicting the inferred origin date of the outbreak using WGS or sites covered by DeepSARS.

### Enabling genomic surveillance of SARS-CoV-2 by identifying regions for targeted deep sequencing

After designing patient- and plate-specific barcodes, we next determined which sites in the viral genome would maximize diagnostic and genomic information while maintaining sufficient coverage for each site. We optimized the sequencing workflow by selecting viral genomic regions ranging from 110 to 140 bp for compatibility with the Illumina MiSeq system (2×81 and 1×150 cycle runs), which ensures a relatively short sequencing duration (<10 hours). The initial viral sites targeted by our protocol were determined by phylogenetic analysis of 2,825 full-length viral genomes from the GISAID repository (December 2019 and March 2020) (Supplementary data file 1). After performing a multiple sequence alignment, we quantified sequence divergence between individual patients at each site in the ∼30,000 base genome of SARS-CoV-2 (Figure 1B), which revealed that certain viral sites contained more mutational diversity than others across the entire genomic landscape (Figure 1B). We next quantified this mutational diversity using sliding regions to uncover the viral sites of 115 bp that maximized the mean sequence divergence across sites between patients. We selected the 13 regions with the most mutational diversity (site-13) and designed corresponding primers that included patientBCs (for nine patients) for evaluation in the deep sequencing library preparation protocol. We furthermore included primers targeting the human RNAse P and human GAPDH genes, which serve as controls and are used in qRT-PCR diagnostics (Vogels et al. 2020; Park et al. 2020).

In light of the recent emergence of variants of concern (VoC), we designed an additional two primer sets that could distinguish the alpha variant (B.1.1.7) across 15 different sites (alpha-15) (Supplementary data file 2) or more generally profile mutational diversity along 16 sites of the spike protein (spike-16) (Supplementary data file 2). Visualizing the primer annealing sites along the full-length viral genome demonstrated the potential to capture sequence diversity for 5,517 bases (18.45%), including the 5’ untranslated region (UTR), ORF1ab, S, ORF3, ORF8 and N genes (Figure 1C).

### Phylogenetic reconstruction of evolutionary and transmission histories via DeepSARS

Since DeepSARS generates sequencing data for targeted regions of SARS-CoV-2, it enables genomic surveillance, which includes inference of evolutionary parameters such as phylogenetic tree topology, mutation rates, and transmission rates (e.g., effective reproductive number, R_e_). We first questioned whether partial sampling (∼20%) of the full-length genome would result in similar tree topologies and variant-specific clustering when compared with WGS. For these benchmarking comparisons, we used data from a recently published study that performed WGS on a localized SARS-CoV-2 outbreak in Austria (Popa et al. 2020). This dataset included viral genomes that were classified according to their geographic location of transmission, thus we determined how using only the targeted genomic regions generated by DeepSARS would impact the tree topology and regional clustering compared to trees inferred from WGS. First, we extracted the targeted sequences (*in silico*) corresponding to site-13, alpha-15 and spike-16 regions from 100 samples containing variant clustering labels as determined by the original authors (Popa et al. 2020); next, we inferred phylogenetic trees rooted by an early SARS-CoV-2 sequence (NCBI reference MN908947.2). This analysis revealed trees generated by the targeted genomic regions of DeepSARS could also lead to variant-specific clustering, although to a lesser extent than trees produced using WGS (Figure 1D).

We next investigated how the targeted sites recovered by DeepSARS could influence the performance of phylodynamic models, which are capable of integrating sampling dates with evolutionary inference. We therefore leveraged Bayesian phylogenetics to infer evolutionary features such the R_e_ value and estimated the origin of the infection on datasets arising from either an early Italian outbreak (before March 2020) or more recently (Feb-Mar 2021); the latter contained a higher proportion of the alpha variant. Supplying either the sites recovered by DeepSARS (combined site-13, alpha-15, and spike-16 primer sets) or WGS as input to a birth-death phylodynamic model revealed comparable distributions of the R_e_ value (Figure 1E). While we observed that combining our three primer sets together resulted in the most comparable R_e_ estimates to WGS, for the alignment containing a high proportion of alpha variants, as expected, the alpha-15 primer set performed especially well (Figures 1E, S2). As we randomly sampled 30 sequences for each of the presented phylodynamic analyses, we resampled new sequences from either time period and assessed how inference using DeepSARS compared to WGS (Figure S2), which again demonstrated highly comparable R_e_ estimations for new sets of sampled sequences. DeepSARS also yielded similar distributions as WGS for estimation of the origin of the outbreak, which is another parameter of infection dynamics (Figures 1F, S2).

### DeepSARS can detect and differentiate viral mutations on synthetic RNA controls of SARS-CoV-2

To evaluate the capacity of DeepSARS for detecting SARS-CoV-2, we performed initial validation using commercially available synthetic RNA controls based on genomic sequences recovered early in the COVID-19 pandemic (NCBI MN908947.3). The initial protocol was performed on a mixture of synthetic viral RNA and human RNA (hRNA), and consisted of RT for addition of patientBCs, followed by PCR with the site-13 pool of primers targeting SARS-CoV-2 and primers for human RNAP and GAPDH. Illumina deep sequencing (1 × 150 bp) resulted in a total of ∼1 million reads, which were then quantified based on containing the correct patientBC and mapping to either the viral genome or human control genes (Figure 2A). When viral RNA and hRNA were present, we were able to selectively amplify and sequence all 13 highly mutational sites, with an average of 781.5 reads per viral site; human gene regions were also detected. In contrast, samples containing either hRNA only or no RNA resulted in minimal background reads (<10 reads) mapping to viral sites, confirming the virus-specific nature of the assay. Next, we determined the limit of detection (LoD) for each viral site by titrating the estimated number of synthetic RNA copies per RT reaction either with or without hRNA. Quantifying the number of mapped reads demonstrated that viral reads could be detected for some sites with as little as 10 copies of synthetic RNA, although the LoD was higher in RT reactions containing hRNA (Figure 2B). Summarizing both the total number of viral reads and the ratio of viral to human reads further revealed the sensitivity to decreasing viral copy numbers. Together, these data support the ability of DeepSARS to simultaneously amplify multiple sites in the SARS-CoV-2 genome in a single reaction at an LoD comparable to diagnostic PCR tests (Vogels et al. 2020).

**Figure 2.**
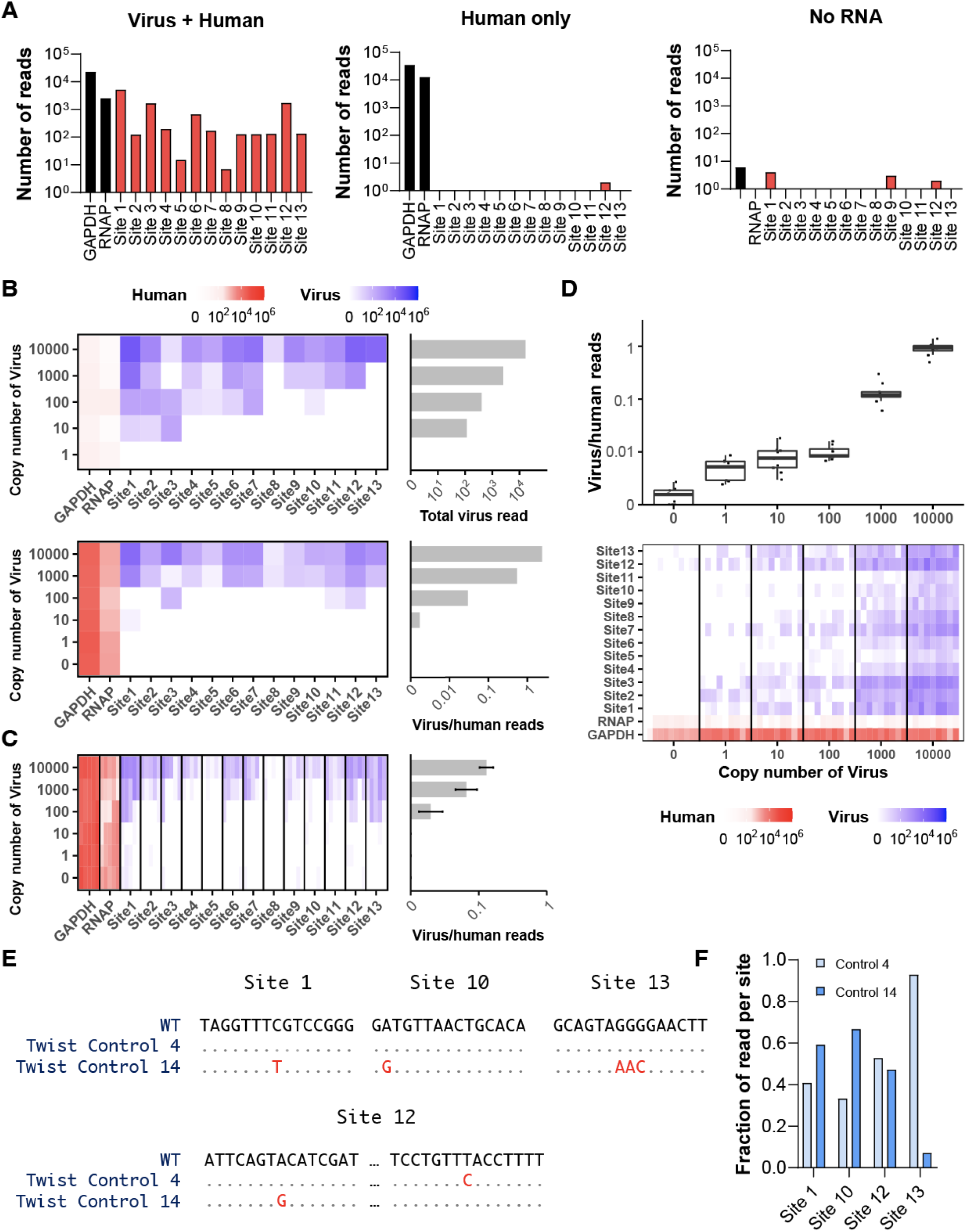
DeepSARS performs highly sensitive detection and identifies mutations from synthetic RNA of SARS-CoV-2. **A**. Samples with synthetic RNA templates of SARS-CoV-2 are mixed with human RNA and subjected to the DeepSARS library preparation protocol using site-13 primer set. Controls include human RNA only and no RNA. Bar graphs show the number of reads mapping to either viral or human genes (GAPDH, RNAP). Each read contained the correct patient barcode. **B**. Synthetic RNA templates are titrated at different copy numbers in the absence (top) and presence (bottom) of human RNA and subjected to the DeepSARS library preparation protocol using site-13 primer set. Heatmaps and bar graphs show reads mapping to viral or human genes and their ratio. **C**. Heatmaps and bar graphs show mapping reads following DeepSARS workflow with site-13 primers on synthetic RNA mixed with human RNA, including pooling samples post reverse transcription (RT). Each column corresponds to a distinct RT reaction. All RT reactions of a given viral copy number used a single patient barcode but a different plate barcode. **D**. Following DeepSARS workflow described as in C but arranged that identical patient barcodes were used for each viral copy number dilution; box plots and heatmaps show the number of viral and human reads. Each column represents an individual RT reaction. Lines separating columns delineate different viral copy number dilutions. **E**. Consensus sequences obtained using the site-13 primer set of DeepSARS on two SARS-CoV-2 synthetic RNA variants with defined mutational diversity. All bases in red indicate expected and recovered mutational divergence. **F**. The fraction of aligned reads containing variant-defining mutations of either synthetic RNA control 4 or synthetic RNA control 14.

In order to further explore the consistency of the DeepSARS library preparation protocol, we performed a number of additional control experiments under various conditions. For example, we pooled six samples arising from distinct RT reactions of different copy numbers of synthetic RNA and hRNA derived from three different cell lines (HEK293, Jurkat, and MCF-7) and performed a multiplex PCR, where each reaction received a single Illumina sequencing adaptor (Figure S3). Following sequencing (Nano MiSeq 1×150 cycles), read alignment, and quantification, we observed individual patientBCs were sensitive to the starting virus copy number, as we used a single patientBC for each copy number dilution (Figure 2C). We next determined if distinct patientBCs recovered reliable numbers of viral reads when using an identical concentration of synthetic viral RNA (Figure S4). Using different patientBCs for each dilution, we performed parallel RT reactions across multiple concentrations of synthetic viral RNA and subsequently pooled nine samples for multiplex PCR, where each reaction received a single Illumina sequencing adaptor. We observed a robust number of reads mapping to a given viral site using different patientBCs, although some sites had more total reads than others (Figure 2D).

After demonstrating the ability of DeepSARS to detect the presence of SARS-CoV-2 with high sensitivity, we next determined whether the recovered sequencing data was sufficient to characterize defined genetic variation between different viral variants. Since the site-13 primers were designed by targeting regions of high genomic diversity, they included four sites (sites 1, 10, 12, 13) that contained point mutations that separated two distinct synthetic RNA control variants (Twist 4 and Twist 14). We therefore performed the DeepSARS workflow and, after sequence read mapping, we could successfully differentiate the two viral control variants (Figure 2E) and their corresponding point mutations with cross-contamination detected in less than 3.8% of reads (Figures 2F, S5).

### DeepSARS enables highly sensitive detection of SARS-CoV-2 from human nasopharyngeal swab and saliva samples

After performing initial validation of DeepSARS on synthetic viral RNA samples, we next assessed its performance on human clinical specimens including nasopharyngeal swabs and saliva. Under standard PCR testing protocols, swab and saliva samples require RNA extraction procedures, most often requiring commercial RNA purification kits. We developed the DeepSARS protocol so that it could be performed on patient samples that were heat-inactivated in guanidinium chloride solution at 95°C, and mixing with 40% Chelex 100 Resin, thereby avoiding the need to explicitly perform RNA extraction with purification kits (Rudolf, Petrillo manuscript in preparation).

We first started with matched samples consisting of six individuals that were previously determined to be COVID-19-positive or -negative based on PCR testing (qRT-PCR). Initially, we performed RT reactions followed by PCR with a distinct combination of both patient- and plate-BCs (unpooled samples during multiplex PCR step). Using the site-13 viral primers and two human gene-specific primers (RNAP and GAPDH), we were able to amplify and recover sequencing reads for the majority of viral sites for both nasopharyngeal and saliva samples from the COVID-19-positive patients (Figure 3A). Alternatively, in all of the COVID-19-negative patients, minimal to no reads were mapped to each viral site (<20 reads total) for both swab and saliva samples (Figure 3A). In order to further validate the sensitivity of DeepSARS, we pooled multiple patient samples (after RT and patientBC addition) within a single PCR (e.g., single plateBC). To this end, we pooled 45 COVID-19-positive and 15 COVID-19-negative patient samples and again observed that DeepSARS could detect significantly higher ratios of viral to hRNA (GAPDH+RNAP) reads in the COVID-19 patient samples (Figures 3B, S6). Importantly, for nasopharyngeal samples, we observed a correlation (R^2^=0.56) between the DeepSARS-generated viral RNA:hRNA read ratio and the cycle threshold (Ct) value as determined by qRT-PCR (Figure 3C). Although DeepSARS consistently recovered fewer reads for saliva samples, we nevertheless observed a correlation between the viral RNA:hRNA ratio and the qRT-PCR Ct values, further supporting the sensitivity of our assay. To better determine technical reproducibility of DeepSARS, we prepared sequencing libraries from both nasopharyngeal and saliva samples for a single individual across multiple patient barcodes and sequencing runs (Patient 35719), which demonstrated a consistent reproducibility regarding the ratio of viral to human reads recovered.

**Figure 3.**
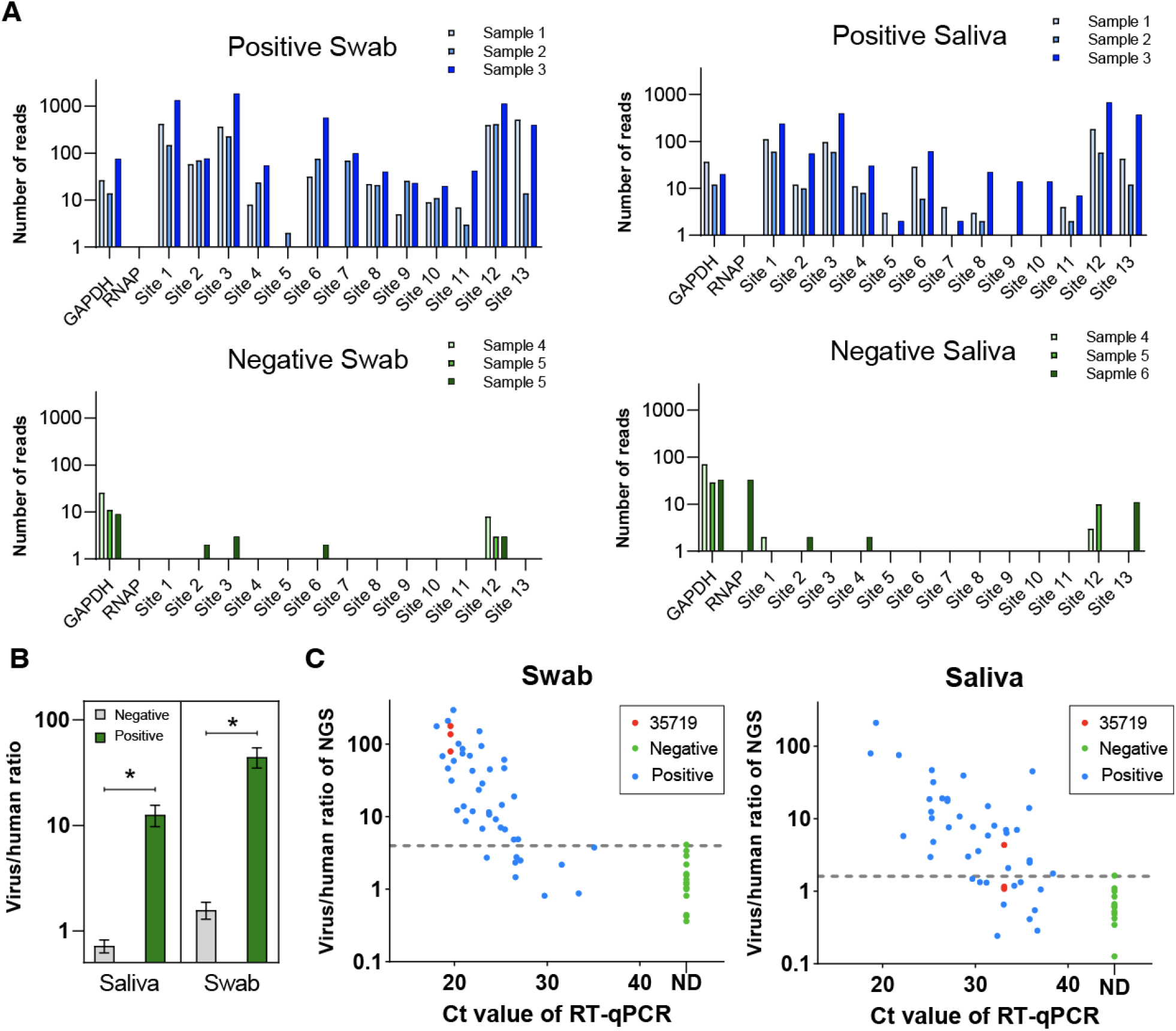
DeepSARS enables diagnostic detection of SARS-CoV-2 from nasopharyngeal and saliva samples. **A**. Nasopharyngeal (swab) and saliva samples of COVID-19 positive and negative patients are subjected to the DeepSARS library preparation protocol using site-13 primer set. Reads mapping to the SARS-CoV-2 genome from three COVID-19 patients and three healthy controls are quantified. Sites 1-13 represent a region in the SARS-CoV-2 genome targeted by the site-13 primer set of DeepSARS. **B**. The ratio of viral to human reads for COVID-19 patients and healthy controls for both swab and saliva samples. Ratio is calculated by summing the reads containing the correct patient barcode across all sites and dividing by the number of reads containing the correct patient barcode mapping to either human GAPDH or human RNAP. **C**. Correlation of CT values determined by qPCR and the ratio of viral to human reads recovered using DeepSARS. Red points (35719) correspond to the same patient included in three independent sequencing runs using different patient barcodes each time.

### Rapid addition of novel viral sites can detect emerging variants of SARS-CoV-2

DeepSARS was initially designed and experimentally validated using 13 sites in the SARS-CoV-2 genome that captured a high genomic sequence divergence at the early stages of the COVID-19 pandemic (April 2020). However, the recent emergence of VoC, such as alpha, beta, gamma, and delta, which have higher transmission rates and are now widespread globally, shows the importance of rapid and diagnostic detection of SARS-CoV-2 variants. We therefore designed and validated the ability of two additional primer sets, one of which covered 15 sites characteristic of the alpha variant (alpha-15) and another targeting 16 sites distributed along the spike protein (spike-16), including several sites present in the RBD (Figure 1C), which possesses mutations that are closely associated with the classification of current VoC. Testing the sets alone on control, or COVID-19 patients with or without the alpha variant demonstrated the ability to recover genomic information for all sites containing the correct patient barcode for both nasopharyngeal and saliva samples using the alpha-15 set (Figures 4A). Combining alpha-15 and site-13 primer sets successfully recovered reads in most sites from COVID-19 samples. Additionally, we were able to selectively pool the primers targeting the spike protein from the site-13 and alpha-15 set with the entire spike-16, again demonstrating the modular ability to mix and match primer sets. Lastly, we combined all three of the primer sets on patient samples, which revealed that, although some sites appeared to be less covered than others, the DeepSARS workflow could be rapidly adapted to handle viral mutations (Figure 4A). Closer inspection into the recovered sequencing reads demonstrated the ability to correctly identify seven mutations characteristic to the alpha variant genome, in addition to detecting patient-specific mutations not found in either the alpha and reference synthetic control (Figure 4B).

**Figure 4.**
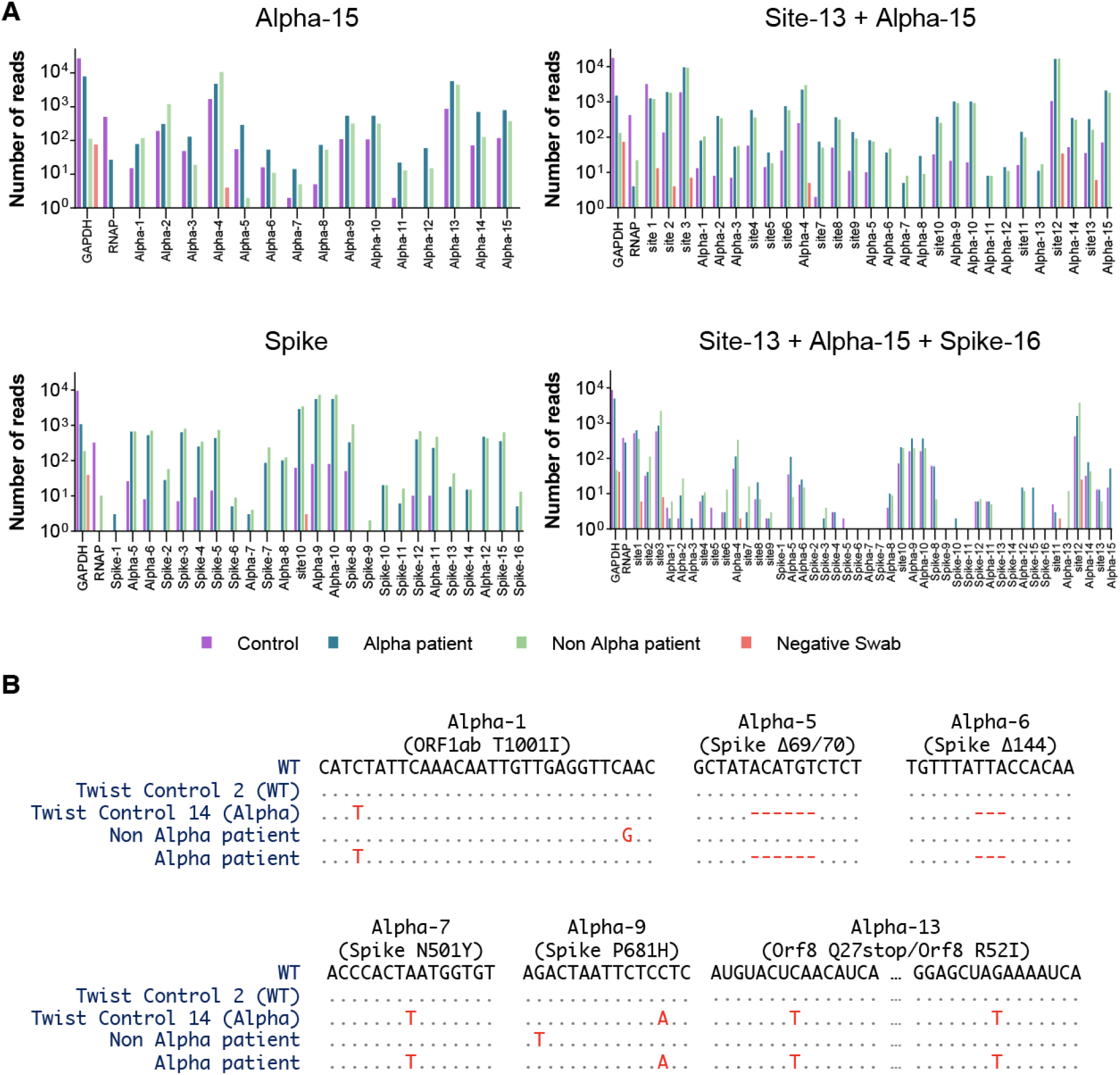
DeepSARS can be rapidly and modularly adapted to perform genomic surveillance by targeting new regions of interest in the SARS-CoV-2 genome. **A**. Nasopharyngeal (swab) samples of COVID-19 positive and negative patients are subjected to the DeepSARS library preparation protocol using primers from site-13, alpha-15, and spike-16 primer sets using one distinct patient barcode and 16 distinct plate barcodes (one per sample per primer set). The number of recovered reads mapping to different regions targeted by DeepSARS on synthetic RNA (control) and three nasopharyngeal patient samples are quantified. Two of the patients had been deemed SARS-CoV-2 positive and one was negative by qPCR. Of the two patients, whole genome sequencing (WGS) had confirmed only one of the two positive cases had the alpha SARS-CoV-2 variant. **B**. Consensus sequences following DeepSARS for the samples in *A* for the sites targeting the mutations defining the alpha SARS-CoV-2 variant. Deletions are indicated as red dashes. Only reads containing the expected patient barcode were included.

### DeepSARS detection of variants over time and across geographic locations

After having experimentally validated the ability of DeepSARS to accurately detect and recover SARS-CoV-2 genomic information from COVID-19 patients, we next determined the extent to which it could be harnessed for classification of VoC and for identifying individual mutations of interest. We first investigated how thoroughly DeepSARS would cover variant-specific mutations located in the spike protein. Cross-referencing the experimentally validated regions revealed that DeepSARS recovered 31 of the 36 defining mutations of alpha, beta, gamma, and delta variants (as determined by US CDC (cdc.gov/coronavirus/2019-ncov/variants/variant-info.html)) (Figure 5A). To confirm that DeepSARS could sufficiently detect population-level dynamics for VoC, we downloaded WGS data from the United Kingdom, South America, and Africa between March 2020 and June 2021 from GISAID (Shu and McCauley 2017). Since the GISAID database provides information regarding the amount and location of mutations, we first quantified the proportion of annotated mutations that we were able to cover using all three of the current DeepSARS primer sets corresponding to the spike region. Each extracted sequence was then classified into viral lineages, including the VoCs, which revealed that DeepSARS was able to detect viral lineage population dynamics at almost the same level as WGS (Figure 5B). Comparing the DeepSARS *in silico* variant classification to the variant annotations for thousands of spike protein sequences sampled at different time points and locations demonstrated the ability to perform genomic surveillance, as the emergence and dominance of certain VoCs in specific locations was accurately identified over time (e.g., alpha and delta in the United Kingdom, gamma in South America, and beta in Africa) (Figure 5B). Quantifying the fraction of annotated spike mutations revealed that DeepSARS could recover sequence information for the majority of annotated amino acid mutations for each full-length spike protein sequence across different locations and time points throughout the pandemic (Figure 5C), highlighting the potential to cover emerging variants and future VoCs. We finally demonstrated that DeepSARS could accurately quantify the dynamic proportion of specific point mutations segregated by geographic location, which was previously profiled using WGS (Scire, Huisman, and Stadler 2021) (Figure 5D).

**Figure 5.**
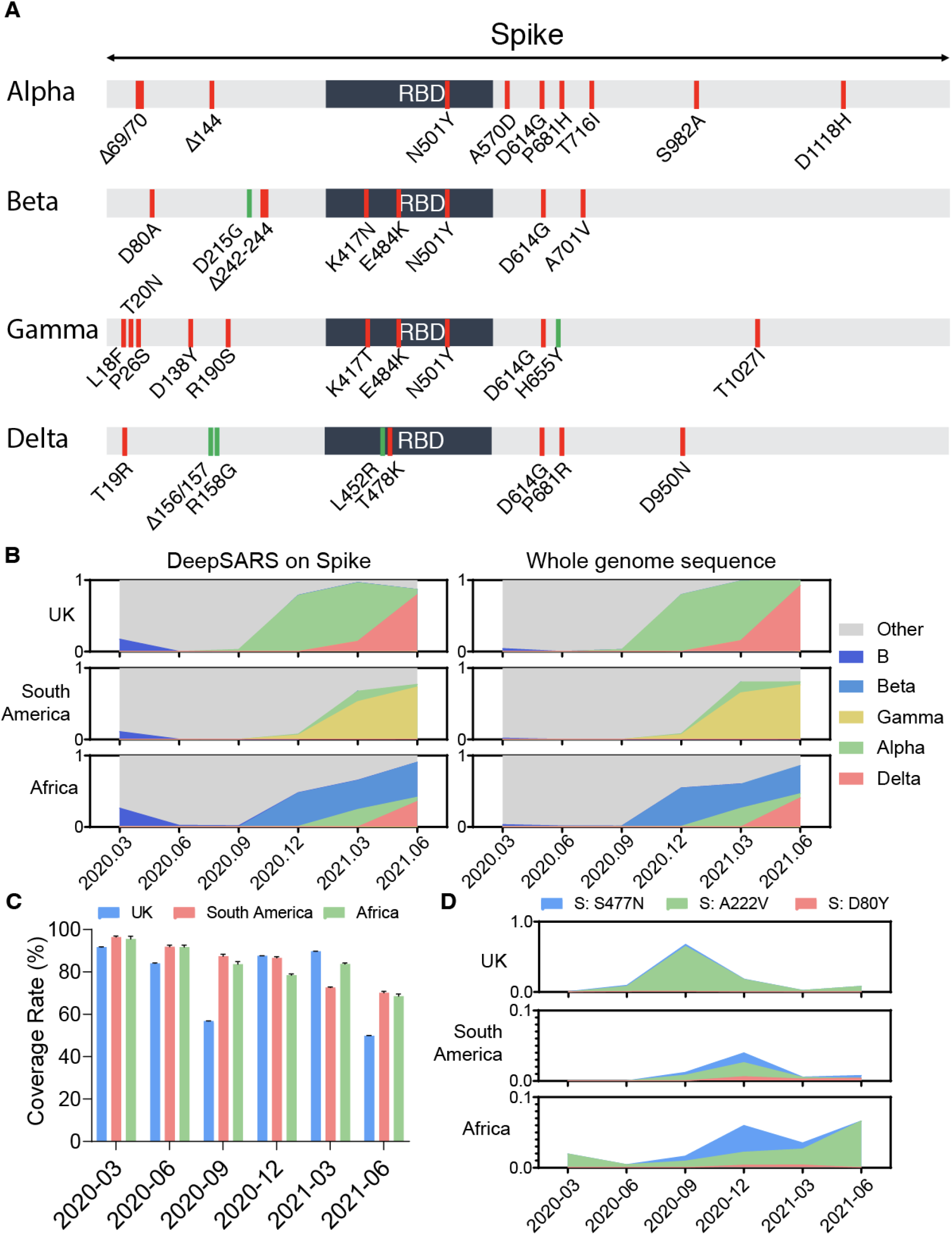
DeepSARS enables variant classification and the quantification of emerging mutations in the spike protein and receptor binding domain (RBD). **A**. Depiction of SARS-CoV-2 spike protein regions targeted by the site-13, alpha-15 and spike-16 primer sets of DeepSARS. Red regions indicate primers present in either the site-13, alpha-15 or spike-16 primer set. Green regions represent those variant-defining mutations not currently targeted by DeepSARS. **B**. Spike-based variant classification for publicly available whole genome sequencing data from GISAID at different time points during the pandemic. Spike protein variants were quantified using either sites covered by DeepSARS or all sites from whole genome sequencing data. **C**. The fraction of mutations covered per spike protein using DeepSARS relative to whole genome sequencing. Error bars indicate standard error of mean. **D**. The fraction of sequences containing one of three previously reported mutations of interest captured by DeepSARS.

## Discussion

Here we have developed DeepSARS, a rapid and scalable approach based on molecular barcoding, targeted deep and computational phylogenetics, thus augmenting basic diagnostic testing with simultaneous genomic surveillance of SARS-CoV-2, which therefore enables detection of VoCs and real-time monitoring of viral evolution and emergence of new variants. We have demonstrated that DeepSARS has excellent sensitivity and specificity on detection SARS-CoV-2 RNA in both extraction-free nasopharyngeal swabs and saliva samples, and correlates with Ct values determined by qRT-PCR (Figure 3C). While this ratio can be used to distinguish the positive and negative samples, it is potentially sensitive to RNA extraction processing, freeze-thaw cycles, and storage conditions. Previous work has also observed a similar phenomenon, where the detection of human RNA was reduced upon storage but the detection of virus RNA was less impacted (Aynaud et al. 2021). Lower human reads may lead to higher DeepSARS-generated viral RNA:hRNA ratio in negative samples, and therefore, could increase false-positives. Future iterations of DeepSARS can increase sensitivity and accuracy by designing other means to classify diagnosis status. While we investigated the relationship between the total number of viral and human reads, in line with previous publications (Ludwig et al. 2021), computational strategies incorporating site-specific information generated by DeepSARS (i.e., how many of the total sites returned reads) or incorporating machine learning methods (i.e., supervised classifier models) may help further improve accuracy and specificity. Moreover, we have demonstrated the possibility to combine all three of the primer sets on patient samples (Figure 4A), although some sites appeared to be less covered than others. This is a common challenge for multiplexing PCR reactions, as different targets in the reaction can compete with each other for resources such that highly abundant templates are preferentially detected, and less abundant ones fade into the background (Elnifro et al. 2000). This can be improved in future iterations by adjusting the ratio of primer for different targets or additional optimization on the PCR condition.

Here we show as a proof-of-concept, DeepSARS performed on a small subset of patient samples, relying on relatively fast but lower read count deep sequencing runs (Illumina MiSeq Reagent Kit v2 Nano, up to one million reads per run). However, DeepSARS can be scaled up and applied to hundreds to thousands of patients in parallel by parallelizing the workflow (number of plates) and increasing deep sequencing depth. For example, ∼4,800 patient samples (∼50 plates) could be prepared by molecular barcoding and sequenced using a single Illumina MiSeq run (2 × 150 kit, 15 million reads). Additional advancements to other sequencing strategies (e.g., NovaSeq with ∼10 billion reads), incorporating automated workflows for library preparation, and using multiple machines in parallel would further improve testing capacity. While the theoretical throughput would be comparable to other targeted deep sequencing-based diagnostics (e.g., SARS-Seq, SwabSeq, C19-SPAR-Seq) (Aynaud et al. 2021; Yelagandula et al. 2021; Bloom et al. 2021), the increased genomic and sequence space coverage provided by DeepSARS enables genomic surveillance. DeepSARS provides genomic surveillance by recovering sequencing information for approximately 20% of the SARS-CoV-2 genome. While DeepSARS resulted in comparable phylodynamic estimates relative to WGS (Figure 1E), we observed that tree topologies diverged from those inferred using WGS (Figure 1D). Accurate estimation of R_e_ will continue to play a critical role in public health policies, as some VoCs have higher R_e_ values and are associated with higher transmission (e.g., alpha and delta variant) (Campbell et al. 2021; Davies et al. 2021). There remains the potential to further optimize the library preparation protocol to allow for increased coverage of the viral genome, which could further improve tree topology estimates. Correctly balancing this trade-off between throughput (multiple patients within a single Illumina sequencing index) and viral coverage (number of targeted genomic regions) holds the potential to balance the inability of qPCR to recover genomic information with the scalability issues of WGS.

A major advantage of DeepSARS is the ability to rapidly adapt the protocol by introducing new primers that cover emerging variants. We have demonstrated three different parameters to which primers could be designed and implemented in DeepSARS, namely by i) mutational divergence from WGS, ii) targeting a specific VoC (e.g., alpha), and iii) targeting viral regions most often associated with VoC (spike protein and RBD). Despite designing and validating these primer sets before the emergence of certain VoCs (e.g., delta and gamma) the viral regions currently recovered by DeepSARS remained relevant and capable of classification of such novel variants (Figures 5A, 5B). Therefore, DeepSARS may enable earlier detection of future variants that may go on to become VoC.

## Methods

### Primer design

Primers based on mutational diversity were designed using 2825 full-length genome sequences (Supplementary data file 1) from the GISAID (April 2020) by calculating the relative number of mutations between whole genome sequences using sliding windows of 150 base pairs. Following identification of viral regions of interest, primers were designed using Geneious (version 10.1.3) to ensure comparable melting temperatures and to avoid disruptive secondary structures. The primer set targeting mutations specific to the alpha (B.1.1.7) variant was designed to first cover the characteristic deletions, followed by those regions in the spike protein, followed by random selection of CDC-classified mutations for remaining regions. The primer set targeting the spike protein prioritized regions within the RBD and then the remaining sites were selected randomly in Geneious, again with comparable melting temperatures and lacking predicted secondary structures.

For the initial reverse transcription step, each patient sample received a patient-specific primer that contains a barcode of at least 10 nucleotides in length. We designed 96 barcodes (one patient barcode per well, 96 wells per plate) that have been computationally optimized to maximize the distance between all barcodes within the set using a genetic algorithm with elitist selection and a random offspring algorithm ensured an average distance of 7.5 mutations for all barcodes, with a minimum distance of 4 mutations between patient barcodes (Figure S1). We additionally tested randomly selected barcodes used in single-cell sequencing (CAGTTGTAGA and AAGTGCCAT) and computationally determined barcodes using edittag (Faircloth and Glenn 2012) and observed no difference in assay sensitivity. Heatmaps displaying barcode distance were performed using the pheatmap R package (Kolde and Kolde 2015).

### Phylogenetic analysis

Maximum likelihood trees were generated using sequences from an early outbreak in Austria for which viral clades had been previously defined (Popa et al. 2020). 100 sequences were randomly sampled from the 418 sequences to better visualize the clustering of clades, as previously performed. Maximum likelihood trees were inferred using the Randomized Axelerated Maximum Likelihood (RAxML) tree construction tool in Geneious (v2020.03) by supplying either the full-length viral genome or just the sites recovered by the three primer sets validated by DeepSARS as input. Trees were then read into R (v.4.0.4) using the “read.tree” function from the R package ape (Paradis, Claude, and Strimmer 2004) and a sequence recovered from Wuhan (MN908947) was set as the outgroup for both trees using the “reroot” function from the phytools R package (Revell 2012).

We assembled five datasets of 30 SARS-CoV-2 genetic sequences with exposure in Italy obtained from publicly available data on GISAID (Shu and McCauley 2017) (accessed on May 2021). For the first two datasets, we randomly selected samples from January 2020 to 8 March 2020 to analyze the early cases documented in Italy. The other three datasets contain samples from February 2021 to 15 March 2021 to describe a later Italian outbreak. We selected sequences for this later outbreak from either i) all lineages, ii) B.1.1.7 lineage (the most frequent lineage at that time in GISAID sequences) and iii) B.1.177 lineage (the second most frequent). Sequences with incomplete collection date, less than 27,000 bases in length or with more than 3,000 unknown bases were omitted. The sequences are aligned with MAFFT under default parameters (Katoh and Standley 2013). The beginning and the end of the alignment are masked respectively by 100 and 50 sequences as well as sites 13402, 24389 and 24390, identified by Nextstrain as prone to sequencing errors (Hadfield et al. 2018). From the sequence alignments, we generate five different alignments by selecting all the sites, the sites included in the primer sets (site-13, alpha-15, or spike-16) and the three primer sets pooled together, thereby producing a total of 25 alignments.

### Phylodynamic analysis

A Bayesian phylodynamic analysis using the BDSKY package (Stadler et al. 2013) of BEAST 2 (Bouckaert et al. 2019) was run for each alignment. The phylogenetic tree was assumed to be produced by a birth-death process with reproductive number R_e_, sampling proportions (s) and becoming uninfectious rate. The sampling proportion for the outbreak was assumed to be zero before the first included sample for that outbreak. We use a LogNormal (0.8, 0.5) prior distribution for R_e_. The value of the becoming uninfectious rate was fixed to 36.5, equivalent to an expected time until becoming uninfectious for each individual of 10 days. We used the HKY substitution model with a strict clock rate fixed to 8×10^−4^ substitutions/site/year (Hadfield et al. 2018). Further, the sampling proportion followed a uniform distribution (0, 0.05), the origin a LogNormal (−2, 0.8), the Kappa (HKY) a LogNormal (1.0, 1.25), the gamma shape (site model) an exponential (0.5), and the gamma category count (site model) was set to 4 (following (Stadler et al. 2013)). We ran one Markov Chain Monte Carlo chain of length 10^7^ iterations for each analysis to approximate the posterior distribution of the model parameters. The chains were assessed for convergence using Tracer v.1.7.1 after discarding the first 10% of samples as burn-in. We ensured that the effective sample size (ESS) was greater than 200 for all parameters. Finally, the posterior probabilities inferred for the parameters of the birth-death process are compared for the alignments with different selected sites for the same set of sequences.

### Patient samples and RT-qPCR

The sample were obtained as part of the project “COVID-19 in Baselland: Validation of Simple and Accurate Tests for COVID-19 Detection, Monitoring and Tracing (ACCURATE-BL-COVID-19)” approved by the ethics board “Ethikkommission Nordwest-und Zentralschweiz (EKNZ)”, Hebelstrasse 53, 4056 Basel representative of Swissethics under the number (2020-01112). Nasopharyngeal swabs and saliva samples were collected, heat-inactivated at 95°C in 3M guanidinium chloride, and subsequently mixed with 40% Chelex 100 Resin at 1:5 ratio. We then assessed 5 μL of patient samples by performing RT-qPCR of a virus specific amplicon (N1) with the GoTaq® Probe 1-Step RT-qPCR System (Promega, A6121) based on methods from CDC.

### DeepSARS sequencing library preparation

RT was performed using 1μL of synthetic RNA control (Twist Biosciences, variants #2, 4, or, 14) or 5 μL of patient samples as a template. Multiplex RT primers (100 μM each) and dNTP (10 mM) were added to the RNA template with a concentration of 5 μM and 0.5 mM, respectively, and incubated for 20 minutes at 70°C. In the case that human RNA was added to synthetic controls, RNA was extracted from HEK293 cell lines and supplied 50 ng. Following incubation, GoScript™ reverse transcriptase and RT buffer (Promega, A5001) were added, incubated at 42°C for 30 minutes followed by 30 minutes at 50°C, and inactivated at 70°C for 15 minutes. For the result shown in Figure 2, Maxima reverse transcriptase (Thermo, EP0742) was used instead. Size-selection was performed using SPRI beads at 1X according to the manufacturer’s protocol to remove RT primers. Where indicated, equal volumes of cDNA were pooled together for a single PCR reaction using a multiplex forward primer set, each of which contained the Illumina TruSeq adaptor sequence and a reverse primer containing an Ilumina index used for demultiplexing (Supplementary data file 2). PCR was performed using KAPA HiFi DNA polymerase (Roche, KK2602) with touchdown PCR. The initial annealing temperature is 67 °C and annealing temperature is gradually reduced (1.1°C/every cycle) for 25 cycles. Amplification is then continued using 58°C for extra 20 cycles. An additional size selection cleanup was performed using SPRI beads at 1.2X according to the manufacturer’s protocol. Library QC was done using a Fragment Analyzer (Agilent Technologies) and libraries were pooled at equimolar concentrations. Final concentration of the pool was measured with the Quant-iT PicoGreen dsDNA Assay Kit (Thermo Fisher Scientific) followed by sequencing using an Illumina MiSeq Reagent Nano Kit v2 (300-cycles) with with an additional 20% PhiX (Illumina) spike-in.

### Bioinformatic analysis and data visualization

A reference genome was created by appending human GAPDH (NM_002046), human RNAP (AL590622), and the full-length reference SARS-CoV-2 genome (MN908947) using Geneious (version 10.1.3). Raw sequencing files were split according to patient specific barcodes allowing for at most one mismatch across the ten nucleotides. Barcode-containing reads were aligned to the reference genome in R using the buildindex function in the Rsubread package (Liao, Smyth, and Shi 2019). Alignment files were exported as bam files and read into R using the loadBam function in the Rsamtools package (Li et al. 2009). Those reads mapping into expected windows based on forward and reverse annealing locations were retained in the analysis. Graphical abstract was created using BioRender.com.

## Data Availability

Data availability: The raw sequencing files that support the findings of this study are available on Zenodo under accession number 10.5281/zenodo.5195784. Code used to design patient barcodes and align and quantify reads can be found at github.com/alexyermanos/deepsars. Additional data that support the findings of this study are available from the corresponding authors upon request.

## Data availability

The raw sequencing files that support the findings of this study are available on Zenodo under accession number 10.5281/zenodo.5195784. Code used to design patient barcodes and align and quantify reads can be found at github.com/alexyermanos/deepsars. Additional data that support the findings of this study are available from the corresponding authors upon request.

## Funding and Acknowledgments

This work was supported by the Botnar Research Centre for Child Health (FTC Covid-19, to S.T.R); Swiss National Science Foundation (Project 31CA30_196348 to STR and TS); European Research Council Starting Grant (Project 679403 to STR) and ETH Zurich Research Grants (to STR and AO).

---

**Figure S1.**
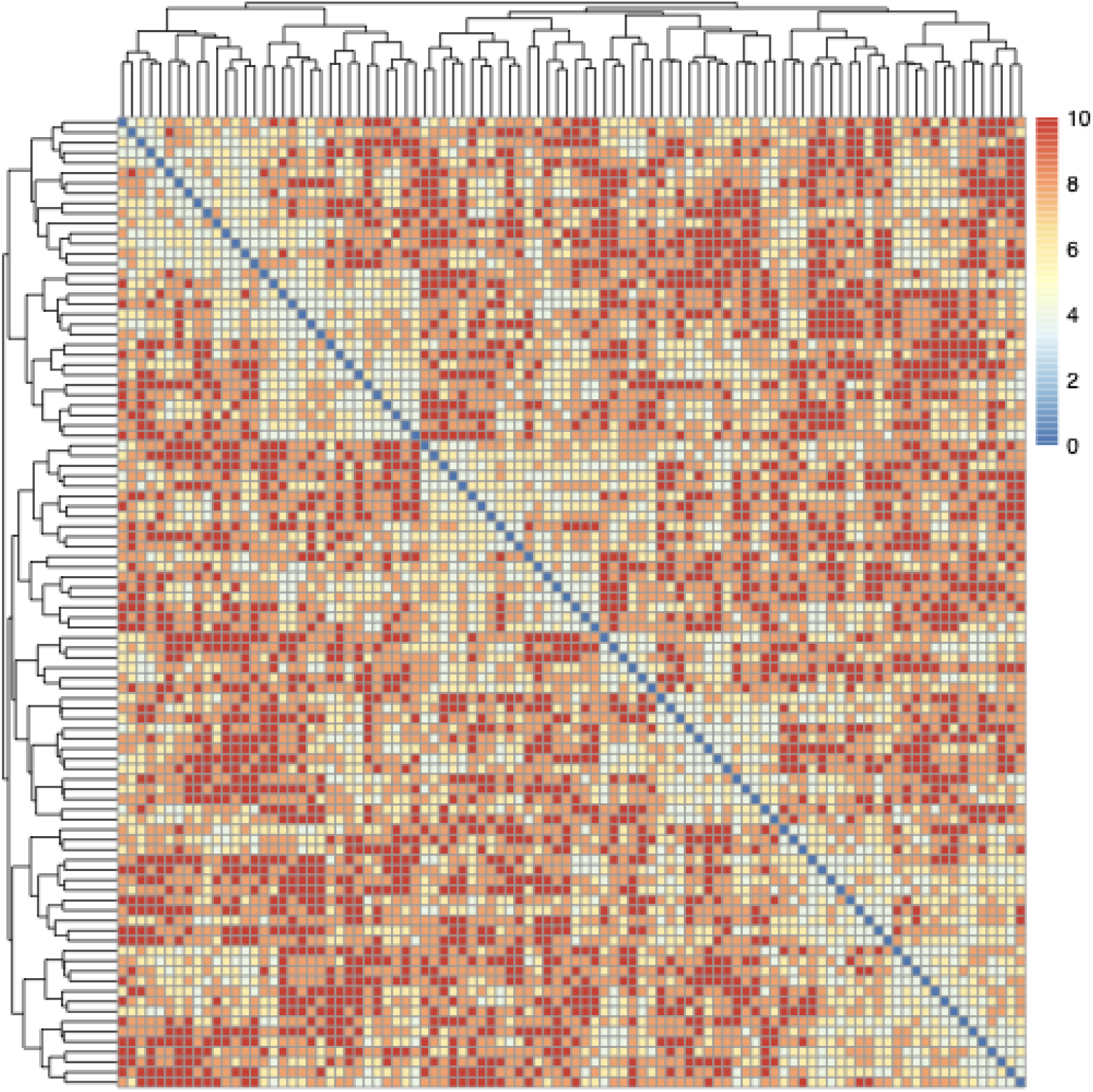
Heatmap demonstrating the number of mutations separating the patient barcodes. The rows/columns correspond to a distinct 10 nucleotide barcode sequence. Intensity corresponds to the number of mutations necessary to convert one barcode into another. Diagonal corresponds to the distance between identical barcodes (distance=0).

**Figure S2.**
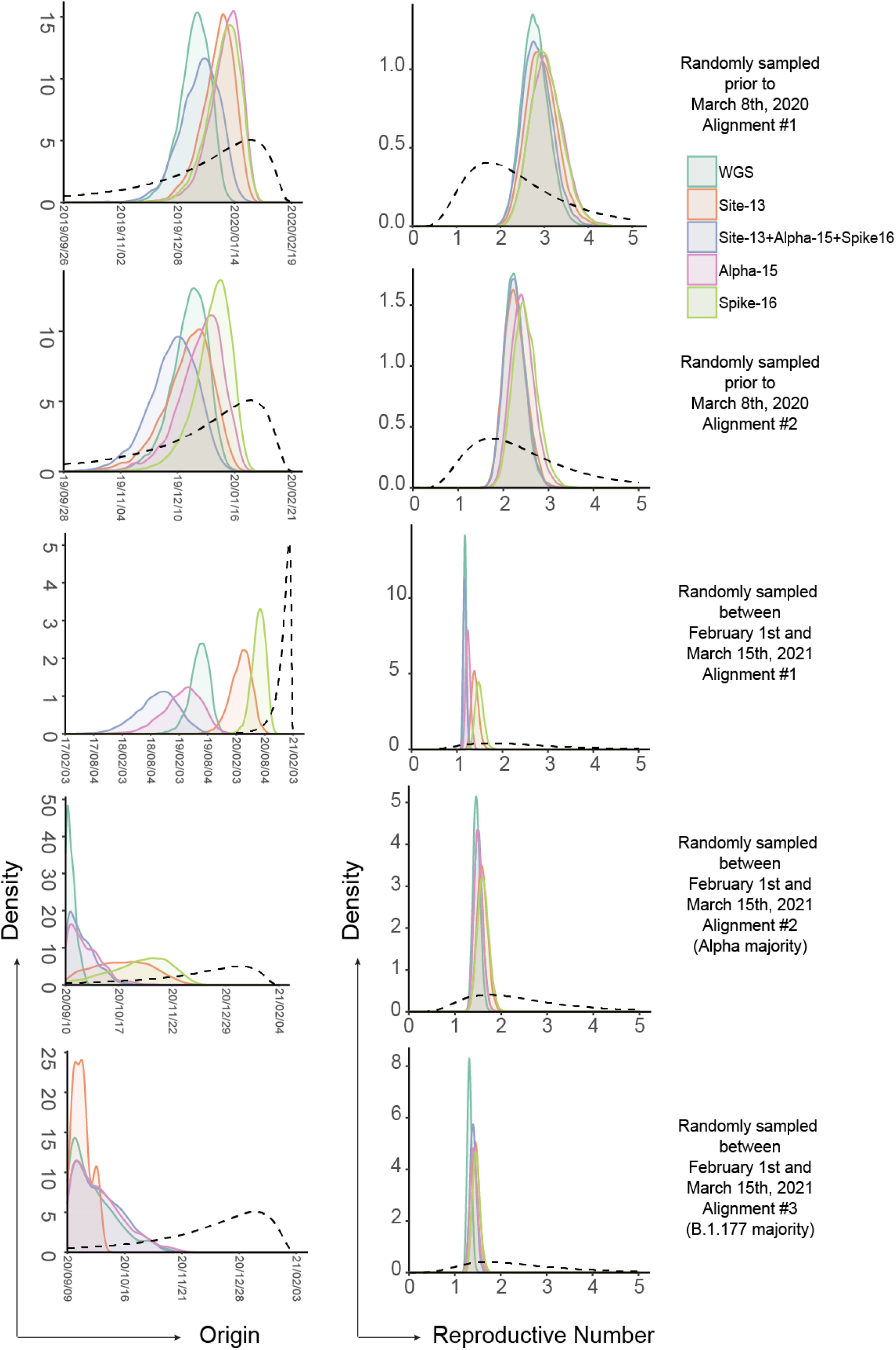
Phylodynamic inference of effective reproduction number and pandemic origin of five different alignments using either sites covered by DeepSARS or all sites in whole genome sequencing. Dashed line indicates the prior distribution.

**Figure S3.**
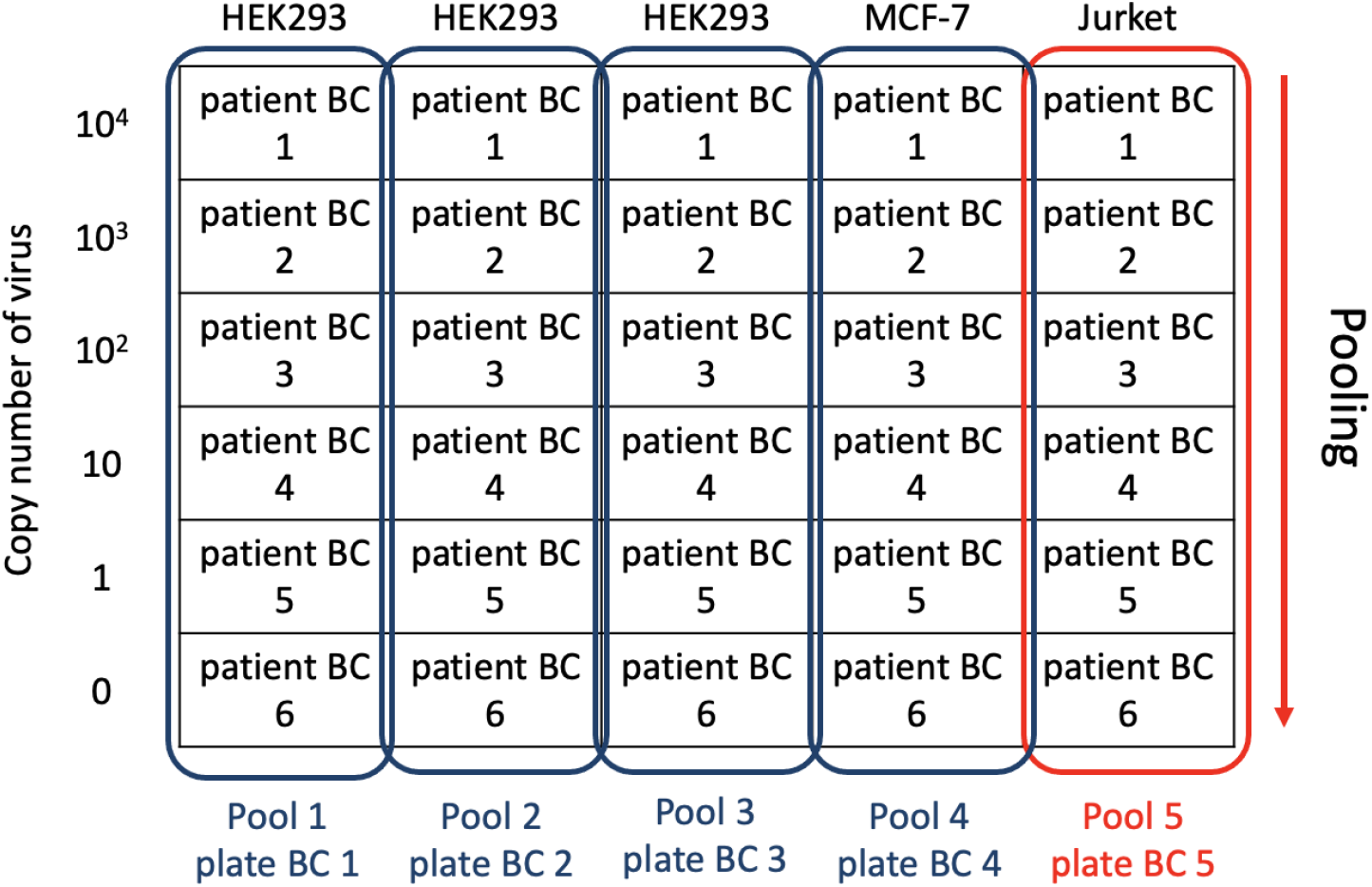
Experimental setting of the experiment depicted in Figure 2C.

**Figure S4.**
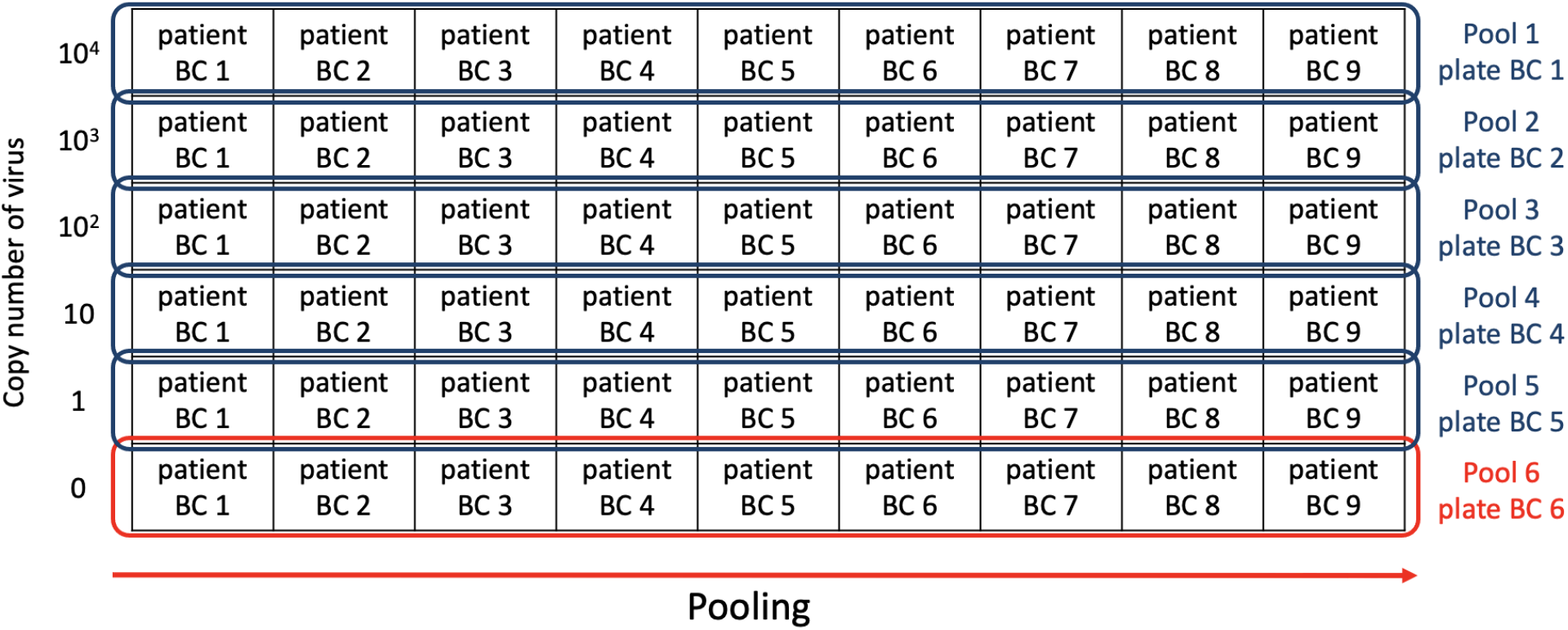
Experimental setting of the experiment depicted in Figure 2D.

**Figure S5.**
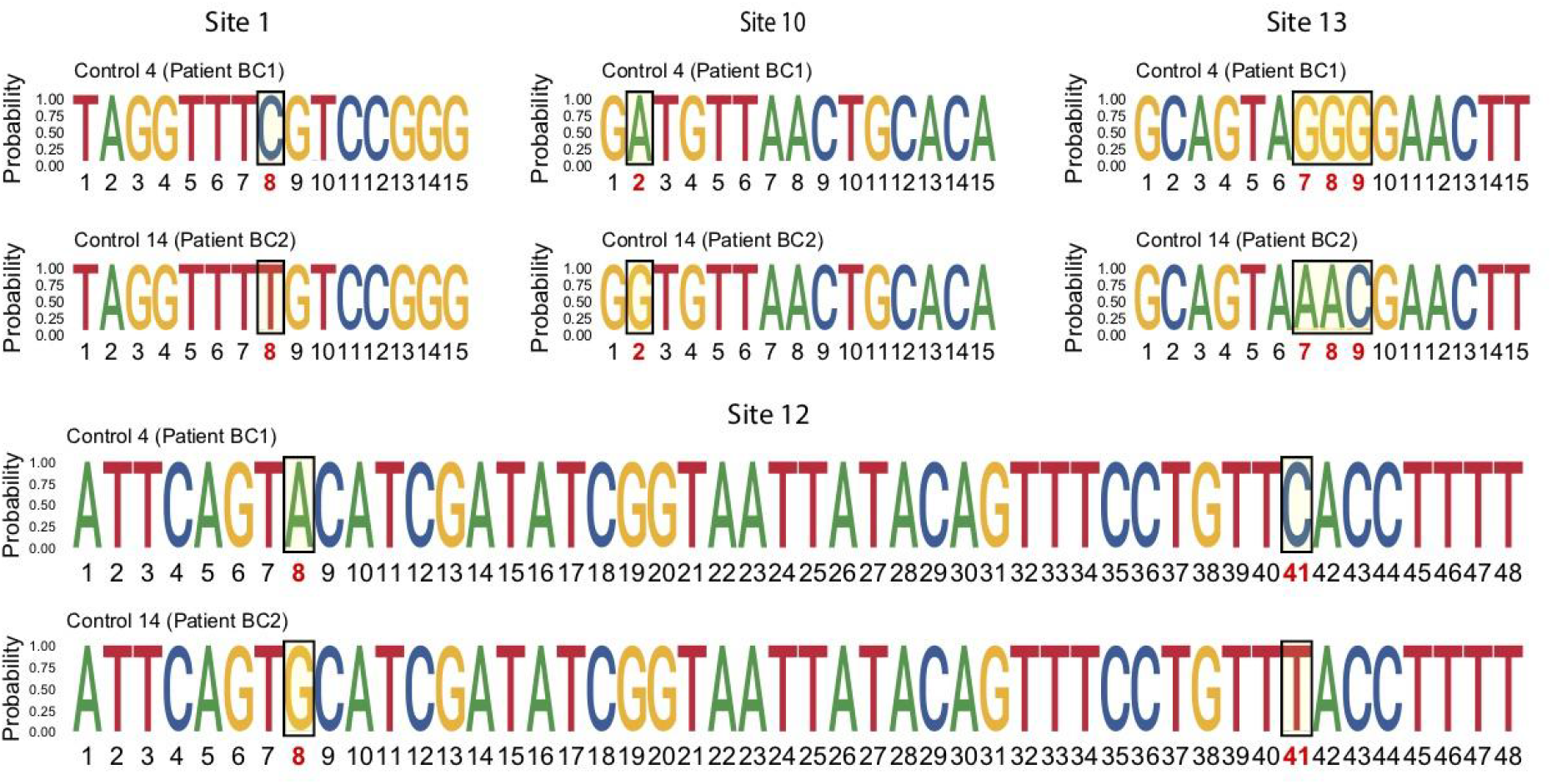
Logo plots depicting the per-base heterogeneity for the **consensus sequences** recovered from all reads in the experiment depicted in Figure 2E. Boxes indicate those sites with defined mutations separating Twist synthetic RNA Controls 4 and 14. Letter size indicates the proportion of reads containing each specific nucleotide at the indicated base.

**Figure S6.**
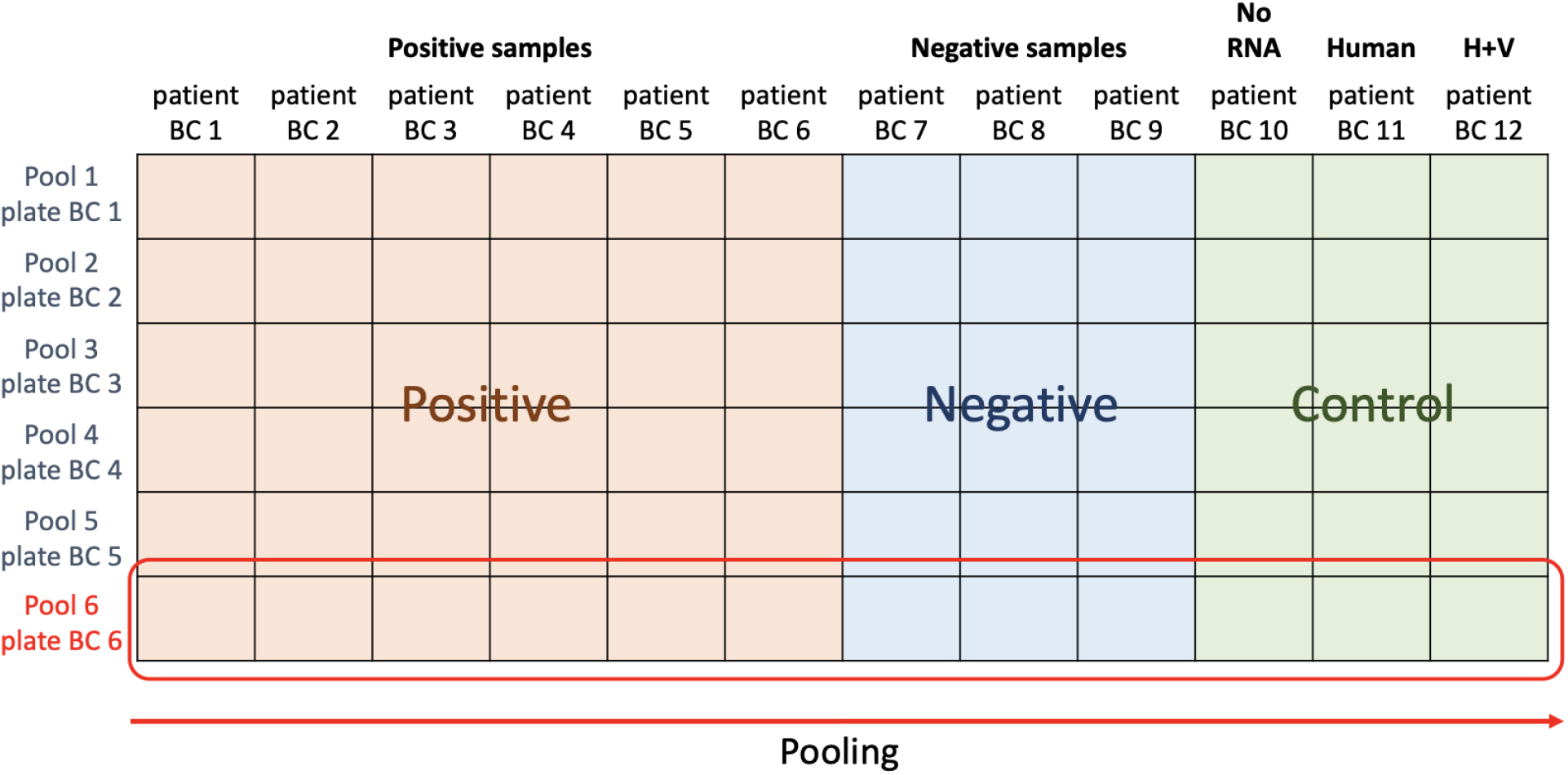
Experimental setting of the experiment depicted in Figure 3.

Supplementary data file 1. Alignment of SARS-CoV-2 genomes used to design site-13 primer set in DeepSARS.

Supplementary data file 2. RT primers and corresponding PCR primers used in DeepSARS.

